# Evaluating a Clinical Decision Support System for Chronic Pain Management: Secondary Per-Protocol Analysis of Pragmatic Randomized Controlled Trial Data

**DOI:** 10.64898/2026.10.07.26364970

**Authors:** Emma McCord, Justin Blackburn, Ann M. Holmes, Nate C. Apathy, Randall W. Grout, Christopher A. Harle, Olena Mazurenko

**Affiliations:** Fay W Boozman College of Public Health, Department of Health Policy and Management, University of Arkansas for Medical Sciences, Little Rock, AR; Indiana University, Indianapolis, IN; Regenstrief Institute, Indianapolis, IN; School of Public Health, Department of Health Policy and Management, The University of Maryland, College Park, MD; Eskenazi Health, Indianapolis, IN, United States

**Author notes:** **Corresponding Author:** Emma McCord, PhD, 4301 W. Markham St., #820-12, Little Rock, AR 72205.

**Keywords:** clinical decision support, chronic pain, electronic health records, pain management, primary care, propensity score matching, per-protocol analysis, implementation science

## Abstract

**Background:** Clinical decision support (CDS) systems may improve chronic pain management by providing guideline-based recommendations within clinical workflows. However, low clinician use can limit their effectiveness. Understanding the association between CDS use and guideline-recommended care may help distinguish implementation challenges from limitations in CDS functionality.

**Objective:** To estimate the association of OneSheet, a CDS system for chronic pain management, and guideline-recommended care when the system was used during patient encounters.

**Methods:** We conducted a per-protocol secondary data analysis of data from a pragmatic randomized clinical trial conducted from October 2020 to May 2022 across 25 primary care clinics at 2 academic health systems in Indiana and North Carolina. The parent trial enrolled 137 primary care clinicians. This analysis included 145511 eligible chronic pain encounters and 15480 long term opioid therapy (LTOT) encounters that occurred after OneSheet implementation. OneSheet use at a patient encounter was the exposure of interest. Propensity score matching was used to create comparable groups of encounters with and without OneSheet use. Outcomes included documentation of pain-related goals, documentation of pain and function using the Pain, Enjoyment of Life and General Activity (PEG) scale, prescription drug monitoring (PDMP) review, urine drug screening (UDS) orders, and naloxone orders.

**Results:** OneSheet was used in 959 of 145511 chronic pain encounters (0.7%) and 582 of 15480 LTOT encounters (4.0%). In matched analyses, OneSheet use was associated with increased documentation of pain-related goals (relative risk (RR)=11.70 (95% CI 4.25-32.50) and PEG score documentation (RR=9.70 (95% CI 4.14-22.31). Among LTOT encounters, OneSheet use was associated with increased UDS ordering (RR=1.52 (95% CI 1.40-1.65) and PDMP review (RR=1.06 (95% CI 1.03-1.09), but decreased naloxone ordering (RR=0.13 (95% CI, 0.02-0.90).

**Conclusions:** OneSheet use was associated with several guideline-recommended chronic pain management actions, suggesting that they system may support care quality when clinicians use it. However, use was rare, indicating that increasing routine use through better workflow integration may be necessary to achieve population-level improvements in chronic pain care.

## Introduction

Clinical decision support (CDS) systems are frequently described as essential for improving care quality, safety, and efficiency [1–3]. By embedding guideline-based recommendations and patient-specific-information into clinical workflows, CDS systems may reduce errors and support informed clinical decision-making. Despite this promise, their real-world impact remains inconsistent [4]. CDS system implementations often face challenges, including alert fatigue, workflow disruptions, usability issues, and clinician skepticism, which limit their effectiveness [5–7]. Subsequently, CDS system use is often low, raising concerns about whether CDS systems can meaningfully influence care quality if clinicians do not actively use them at a patient encounter [4–8].

Variability in CDS system impact is often attributed to differences in actual clinician use [12]. CDS systems can only influence care when clinicians meaningfully engage with them.^13-16^ For example, poor alignment with clinical workflow can lead to low use, undermining the potential benefits of the CDS system [16–19]. For this reason, evaluations based solely on intention-to-treat (ITT) analyses may underestimate the potential impact of CDS systems when use is low because these analyses assess the effect of access rather than engagement [20–21]. In contrast, per-protocol analyses focus on encounters in which clinicians use the CDS system and therefore provide insight into its potential impact under successful implementation [20–21]. Consequently, per-protocol analyses can help distinguish whether limited improvements are primarily attributable to low use or to limitations in CDS functionality itself.

Given the time constraints and complexity of patients’ pain histories, CDS systems for chronic pain are a prime candidate for helping primary care clinicians (PCCs) deliver guideline-recommended pain care [22–24]. One such CDS system for chronic pain care, OneSheet, was implemented as part of a pragmatic randomized controlled trial (RCT) across two academic health systems and 25 clinics [18,19,25,26]. OneSheet presents guideline relevant information (e.g., pain diagnoses, pain and function goals) in a single electronic health record (EHR) view and provides shortcuts to recommended clinical actions (e.g., order urine drug screen (UDS)) [25]. Several prior publications have reported different aspects of OneSheet, including its user-centered design and usability testing, qualitative evaluations of barriers and facilitators to use, and the parent ITT analysis evaluating access to OneSheet [18,19,25,26,27]. The current study uses the same trial population, encounter data, and outcome definitions as the parent trial but addresses a distinct question. Whereas the parent ITT analysis estimated the effect of providing PCCs access to OneSheet on guideline-recommend chronic pain care [27], the current per-protocol analysis examines the associations between actual encounter-level OneSheet use and guideline-recommended care. Thus, the purpose this study was to estimate the association of OneSheet with guideline-recommended chronic pain care when it was used at a patient encounter.

## Methods

### Study Design and Setting

We analyzed secondary data from a pragmatic RCT conducted from October 2020 to May 2022 across two academic health systems, one in Indiana and one in North Carolina, USA. The trial enrolled 137 PCCs from 25 clinics; 69 PCCs were randomized to the intervention group and received access to OneSheet in their Epic EHRs. PCCs were eligible to enroll if they were actively practicing at participating clinics, licensed to prescribe medications, and trained in family medicine or related specialties. Recruitment occurred in two phases during 2020 and 2021. Additional details of our recruitment strategy and trial design are reported elsewhere [25–28].

For the per-protocol analysis, the unit of analysis was the patient encounter, nested within patients and PCCs. We restricted the analytic sample to encounters with patients who qualified as having chronic pain, identified using a pre-defined algorithm (**see Figure 1**). Because three of our outcome variables are only relevant for encounters in which the patient was on long-term opioid therapy (LTOT), we also identified the subset of this sample in which the patient was prescribed at least a 28-day supply of an opioid analgesic at the time of the encounter [22]. Figure 2 illustrates how the analytic groups were derived from the original RCT data. Because our focus was on the association of OneSheet when it was available, both groups were restricted to encounters occurring after OneSheet implementation.

**Figure 1.**
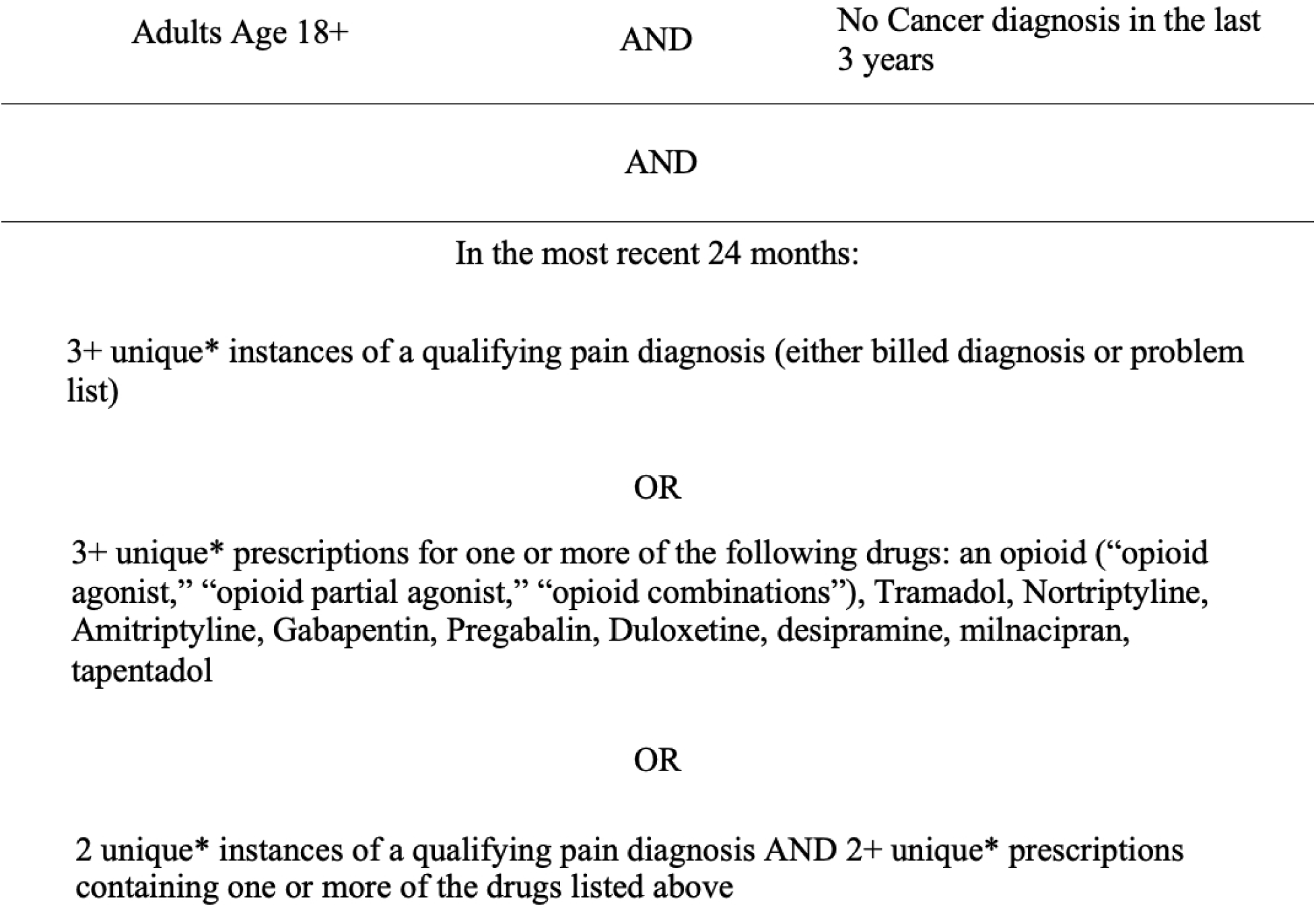
Approach for Identifying Patients with Chronic Pain.

**Figure 2.**
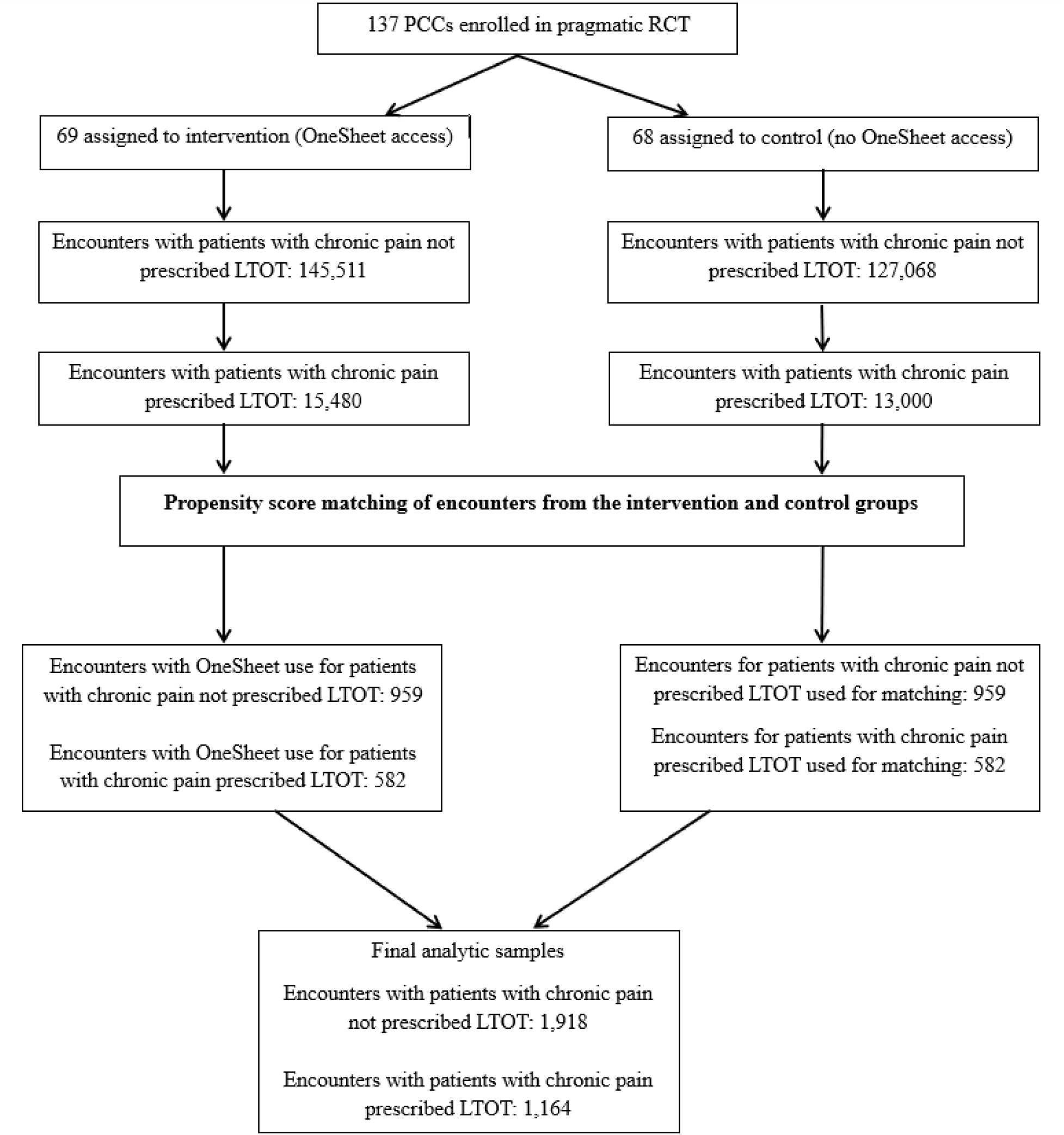
Approach for Identifying Chronic Pain Encounters Using Propensity Score Matching.

### OneSheet Exposure

We defined OneSheet use at a patient encounter using EHR usage data containing unique PCC identifiers and corresponding OneSheet access dates. Because usage data provided for this study recorded access dates rather than direct encounter identifiers, each OneSheet access was attributed to the nearest encounter for the corresponding patient-PCC pair. To minimize misclassification, each access event was assigned to only one encounter and was retained only when the nearest encounter occurred on or within three days of the access date. This approach was selected based on the empirical distribution of encounter dates relative to access dates and prior qualitative evidence indicating how PCCs commonly used OneSheet when preparing for upcoming visits and when completing follow-up tasks after encounters [18]. Encounters occurring outside the 7-day window were considered unrelated to the observed OneSheet access.

### Outcome Measures

Using EHR data, we constructed five encounter-level binary outcomes that reflected concordance with the 2016 Centers for Disease Control and Prevention (CDC) opioid prescribing guideline recommendations (1 = guideline-concordant, 0 = non-concordance) [23]. For all encounters with patients identified as having chronic pain, the outcomes included: (1) documenting pain-related goals in the last 90 days and (2) documenting pain and function via Pain, Enjoyment of Life and General Activity (PEG) scale in the last 90 days. For the smaller subset of encounters in which patients were prescribed LTOT, the outcomes included: (3) reviewing prescription drug monitoring program (PDMP) in the last 90 days; (4) ordering UDS results in the last calendar year; (5) and ordering naloxone within 7 days of the encounter in which an opioid was prescribed. Consistent with the parent trial, these outcomes were intended to measure guideline-concordant care status rather than clinical actions occurring exclusively during the indexed encounter. Thus, encounters could be considered concordant when activities had occurred within the clinically relevant lookback period. A comprehensive explanation of our outcome construction is provided elsewhere [20].

### Propensity Score Matching

OneSheet was used in fewer than one percent of encounters in the sample randomized to treatment. Because of this differential missingness, randomization failed to produce balance in the observable characteristics in the treatment and control arms for a per-protocol analysis. Thus, propensity score models were used to minimize biases caused by baseline differences to match treatment and control encounters on their propensity to use OneSheet.

Two separate propensity score models were estimated. In the first, the analysis was restricted to all encounters in which the patient had chronic pain and occurred after OneSheet was implemented. Logistic regression was then applied to the sample of encounters randomized to treatment to identify observable encounter characteristics associated with the use of OneSheet. Covariates included patient demographics (i.e., gender, race, ethnicity, age), PCC characteristics (i.e., gender, experience, clinic site), and encounter-level factors (i.e., whether a chronic pain diagnosis was attached to the encounter, whether the patient was new to the PCC). The resulting parameter estimates were then applied to the sample of encounters randomized to the control group to generate predicted probabilities of OneSheet use (i.e., propensity scores). Optimal 1:1 matching was performed using the MatchIt package in R (version 4.7.2) to pair encounters in which OneSheet was used to comparable encounters from the control group [32]. This matched sample was appropriate for evaluating documentation of pain-related goals and PEG scores which are applicable for all encounters in which the patient has chronic pain. This approach improves the comparability of matched samples and reduces bias, particularly when overlap between groups is imperfect [29,30,33]. Guidelines only recommend PDMP review, UDS orders, and naloxone orders for patients on LTOT. For this reason, we performed a second propensity score exercise limited to the sample of encounters in which the patient had chronic pain and was on LTOT as described above, but removing LTOT from the list of covariates included in the logistic regression. This second matched sample was used to evaluate OneSheet use on PDMP review, UDS orders, and naloxone orders. Acceptable covariate balance was achieved across all measured characteristics using matching without replacement and without any caliper restrictions.

After matching, we evaluated the covariate balance between encounters with OneSheet use and their matched control encounters to ensure that propensity score matching effectively reduced baseline differences. Standardized mean differences (SMDs) were calculated for all covariates before and after matching, with a SMD < 0.1 indicating acceptable balance. We also performed visual checks, including propensity score distribution overlaps and standardized difference plots, to confirm sufficient overlap between the two groups.

### Statistical Analysis

After matching, we estimated the association between OneSheet use at patient encounters and our five binary outcomes. For each outcome, we calculated the baseline risk (e.g., the proportion of control encounters without OneSheet use in which the guideline-recommended action was observed). We also estimated relative risks (RR) by exponentiating coefficients from modified Poisson regression models with robust standard errors. Robust standard errors were clustered at the PCC level to account for correlation of encounters managed by the same PCC. RR represents the ratio of the probability of the outcome occurring at encounters with OneSheet use to the probability of the outcome occurring at control encounters without OneSheet use. Values above 1 indicate a higher likelihood with OneSheet use, whereas values below 1 indicate a lower likelihood. We report 95% confidence intervals and p-values from these models, with p-values < 0.05 considered statistically significant. This method provides directly interpretable risks and accounts for matched samples using robust variance estimation [34]. We also describe differences between control encounters and encounters with OneSheet use in terms of absolute risk difference. The absolute risk difference is the percentage of encounters with OneSheet use in which the guideline-recommended action was observed compared to the percentage of control encounters without OneSheet use in which the guideline-recommended action was observed. To visually summarize the results, we created forest plots. All analyses were performed in R (version 4.4.1).

### Ethical Considerations

The study complied with the World Medical Association Declaration of Helsinki on Ethical Principles for Medical Research Involving Human Subjects. The Indiana University Institutional Review Board approved the study protocol. Study participants (PCCs) signed an informed consent. A waiver of patient consent was approved because patient data was routinely collected in the EHR and a high volume of patient visits, making obtaining informed consent not feasible.

## Results

### Study Sample

Among intervention PCCs with access to OneSheet, 145511 eligible chronic pain encounters and 15480 eligible LTOT encounters occurred during the study period (**see Figure 2**). OneSheet was used during 959 (0.7%) chronic pain encounters and 582 (4%) LTOT encounters. Following propensity score matching, the final analytic sample included 1,918 chronic pain encounters and 1164 LTOT encounters. Characteristics of chronic pain and LTOT encounters are presented in Tables 1 and 2, respectively.

**Table 1.** Comparison of Measurable Characteristics of the Matched Sample of Chronic Pain Patients.

| Characteristic | Used OneSheet at encounter |  | SMD |
| --- | --- | --- | --- |
|  | Yes (n=959) | No (n=959) |  |
| Patient characteristics |  |  |  |
| *Gender* |  |  |  |
| Female | 0.65 | 0.65 | <0.01 |
| Male | 0.35 | 0.35 | <0.01 |
| *Race* |  |  |  |
| White or Caucasian | 0.31 | 0.31 | <0.01 |
| Black or African American | 0.67 | 0.67 | <0.01 |
| Other | 0.02 | 0.02 | <0.01 |
| *Ethnicity* |  |  |  |
| Not Hispanic or Latino | 0.99 | 0.98 | <0.01 |
| Hispanic or Latino | 0.01 | 0.02 | 0.01 |
| Unknown | 0.00 | 0.00 | <0.01 |
| *Age* |  |  |  |
| 0-47 years old | 0.12 | 0.11 | 0.01 |
| 48-56 years old | 0.23 | 0.24 | 0.01 |
| 57-63 years old | 0.22 | 0.22 | 0.01 |
| 64-70 years old | 0.27 | 0.27 | 0.01 |
| 71+ years old | 0.16 | 0.16 | <0.01 |
| PCC characteristics |  |  |  |
| *Gender* |  |  |  |
| Female | 0.84 | 0.85 | 0.01 |
| Male | 0.16 | 0.15 | 0.01 |
| *Experience* |  |  |  |
| 0-13 years | 0.18 | 0.18 | <0.01 |
| 14+ years | 0.82 | 0.82 | <0.01 |
| *Site* |  |  |  |
| System #1 | 0.85 | 0.84 | <0.01 |
| System #2 | 0.15 | 0.16 | <0.01 |
| Encounter characteristics |  |  |  |
| *Chronic pain diagnosis attached to encounter* |  |  |  |
| Yes | 0.73 | 0.72 | 0.01 |
| No | 0.27 | 0.28 | 0.01 |
| *Patient is new to PCC* |  |  |  |
| Yes | 0.02 | 0.03 | 0.1 |
| No | 0.98 | 0.97 | 0.1 |
| *Patient is prescribed LTOT* |  |  |  |
| Yes | 0.61 | 0.61 | <0.01 |
| No | 0.39 | 0.39 | <0.01 |

**Table 2.**
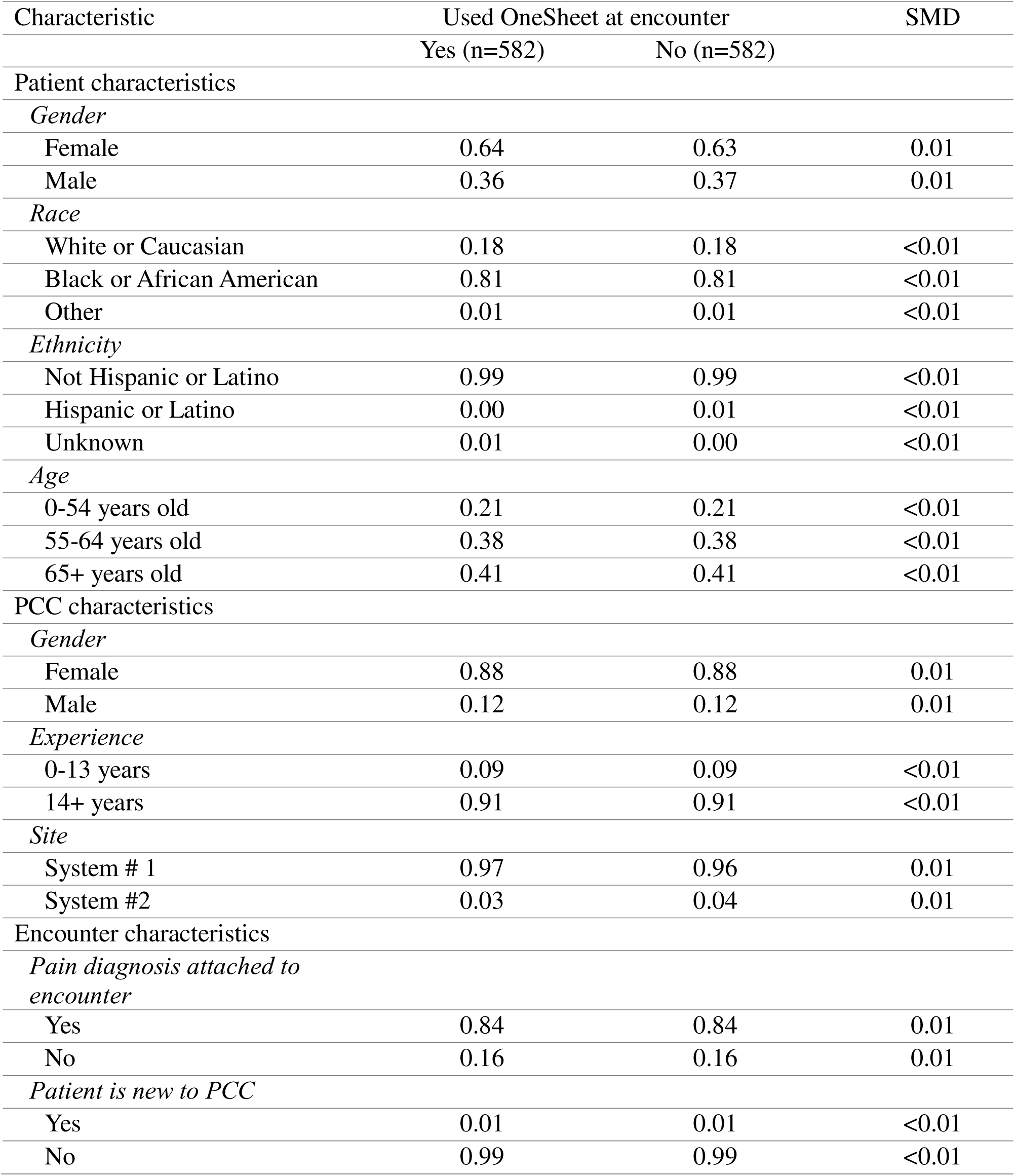
Comparison of Measurable Characteristics of the Matched Sample of Patients Prescribed LTOT.

| Characteristic | Used OneSheet at encounter |  | SMD |
| --- | --- | --- | --- |
|  | Yes (n=582) | No (n=582) |  |
| Patient characteristics |  |  |  |
| *Gender* |  |  |  |
| Female | 0.64 | 0.63 | 0.01 |
| Male | 0.36 | 0.37 | 0.01 |
| *Race* |  |  |  |
| White or Caucasian | 0.18 | 0.18 | <0.01 |
| Black or African American | 0.81 | 0.81 | <0.01 |
| Other | 0.01 | 0.01 | <0.01 |
| *Ethnicity* |  |  |  |
| Not Hispanic or Latino | 0.99 | 0.99 | <0.01 |
| Hispanic or Latino | 0.00 | 0.01 | <0.01 |
| Unknown | 0.01 | 0.00 | <0.01 |
| *Age* |  |  |  |
| 0-54 years old | 0.21 | 0.21 | <0.01 |
| 55-64 years old | 0.38 | 0.38 | <0.01 |
| 65+ years old | 0.41 | 0.41 | <0.01 |
| PCC characteristics |  |  |  |
| *Gender* |  |  |  |
| Female | 0.88 | 0.88 | 0.01 |
| Male | 0.12 | 0.12 | 0.01 |
| *Experience* |  |  |  |
| 0-13 years | 0.09 | 0.09 | <0.01 |
| 14+ years | 0.91 | 0.91 | <0.01 |
| *Site* |  |  |  |
| System # 1 | 0.97 | 0.96 | 0.01 |
| System #2 | 0.03 | 0.04 | 0.01 |
| Encounter characteristics |  |  |  |
| *Pain diagnosis attached to encounter* |  |  |  |
| Yes | 0.84 | 0.84 | 0.01 |
| No | 0.16 | 0.16 | 0.01 |
| *Patient is new to PCC* |  |  |  |
| Yes | 0.01 | 0.01 | <0.01 |
| No | 0.99 | 0.99 | <0.01 |

### Propensity Score Matching and Covariate Balance

In both the full sample of patients with chronic pain and the subsample of patients prescribed LTOT, matching reduced standardized differences between encounters with and without OneSheet use to below 0.1 for all covariates, indicating good balance [35].

### Outcomes Among Chronic Pain Encounters

In the matched sample of chronic pain encounters, documentation of pain-related goals occurred in 0.4% of matched encounters randomized to control compared with 4.7% of matched encounters randomized to treatment in which OneSheet was used. Thus, encounters with OneSheet use were 11.7 times more likely to include documentation of pain-related goals than matched control encounters without OneSheet use (RR=11.70 (95% Confidence Interval (CI) 4.25-32.50), with an absolute risk difference of 4.3% (see Table 3). Documentation of PEG scores occurred in 1.0% of matched control encounters compared with 9.7% of matched encounters with OneSheet use. Thus, encounters with OneSheet use were 9.7 times more likely to include documentation of PEG scores than matched control encounters without OneSheet use (RR=9.70 (95% CI 4.19-22.31), with an absolute risk difference of 9.7%.

**Table 3.** Association Between OneSheet Use at a Patient Encounter and Guideline-Recommended Chronic Pain Management (Per-protocol Analysis)

| Sample | OneSheet users | Matched sample of non-users<sup>a</sup> | Calculated RR of user to non-user<sup>b</sup> | 95% CI |
| --- | --- | --- | --- | --- |
| Percent achieved |  |  |  |  |
| Document pain-related goals<sup>c</sup> | 4.70 | 0.40 | 11.70<sup>h</sup> | (4.25-32.50) |
| Document PEG<sup>d</sup> | 9.70 | 1.00 | 9.70<sup>h</sup> | (4.19-22.31) |
| Order UDS<sup>e</sup> | 82.00 | 54.00 | 1.52<sup>h</sup> | (1.40-1.65) |
| Review PDMP<sup>f</sup> | 98.00 | 93.00 | 1.06<sup>h</sup> | (1.03-1.09) |
| Order Naloxone<sup>g</sup> | 0.10 | 1.00 | 0.13<sup>i</sup> | (0.02-0.90) |
| Sample size (full chronic pain n=1918; LTOT n=1164) |  |  |  |  |

<sup>a</sup>Baseline risk was the proportion of control encounters without OneSheet use in which the outcome of interest could be observed.
<sup>b</sup>Relative effects/risks are the exponentiated coefficients that were estimated using modified Poisson regression with robust standard errors. All analyses are based on matched samples (1:1 optimal propensity score matching).
<sup>c</sup>Goals- documenting pain-related goals in the last 90 days.
<sup>d</sup>PEG- documenting pain and function via Pain, Enjoyment of Life and General Activity (PEG) scale in the last 90 days.
<sup>e</sup>PDMP- reviewing prescription drug monitoring program in the last 90 days.
<sup>f</sup>UDS- ordering urine drug screen. results in the last calendar year.
<sup>g</sup>Naloxone- ordering Naloxone within 7 days of the visit.
<sup>h</sup>p<0.05.
<sup>i</sup>p<0.001.

### Outcomes Among LTOT Encounters

Among matched encounters involving patients prescribed LTOT, UDS was ordered in 54% of matched encounters randomized to control and in 82% of matched encounters randomized to treatment in which OneSheet was used. Thus, encounters with OneSheet use were 1.52 times more likely to include a UDS order than matched control encounters without OneSheet use (RR=1.52 (95% CI 1.40-1.65), with an absolute risk difference of 28% (see Table 3). PDMP review occurred in 93% of matched control encounters and 98% of matched encounters with OneSheet use. Thus, encounters with OneSheet use were 1.06 times more likely to include a PDMP review than matched control encounters without OneSheet use (encounter (RR=1.06 (95% CI 1.03-1.09), with an absolute risk difference of 5%. Finally, naloxone was ordered in 1.0% of matched control encounters and 0.1% of matched encounters with OneSheet use. Thus, encounters with OneSheet use were 0.87 times less likely to order naloxone than matched control encounters without OneSheet use (RR=0.13 (95% CI 0.02-0.90), with an absolute risk difference of -0.09%.

## Discussion

### Principal Findings

Our per-protocol analysis suggests that OneSheet use was associated with improvements in several guideline-recommended chronic pain actions. Encounters in which OneSheet was used were more likely to include documentation of pain-related goals and PEG scores and more likely to include UDS orders and PDMP review compared with matched control encounters. These findings indicate that OneSheet may support guideline-recommended care when clinicians engage with the system.

OneSheet was used in only 0.7% of chronic pain encounters and 4.0% of LTOT encounters. Although the observed associations suggest that OneSheet may support guideline-recommended care, the low rate of use indicates that these benefits would be difficult to achieve at a population level without more consistent engagement. These findings highlight the importance of integrating CDS systems into routine clinical workflows in ways that promote sustained use.

Encounters with OneSheet use were more likely to document pain-related goals and PEG scores compared with matched control encounters. Although the relative increases were substantial, absolute rates remained low. Even among encounters in which OneSheet was used, pain-related goal documentation occurred in only 4.7% of encounters and PEG documentation occurred in 9.7%. These findings suggest that barriers beyond information access likely limit routine documentation of these patient-centered measures. Moreover, neither local nor national policies mandate clinicians to document these outcomes, yet they are emphasized in the CDC opioid prescribing guidelines as essential to patient-centered care [22–24]. To date, they remain rarely and inconsistently documented in primary care [8,36–42]. Only a small number of CDS systems offer structured features to support these practices [13], but our findings show that when these features are available in a CDS system that PCCs use, adoption of recommended practices increases. Our findings suggest that CDS systems can help address this gap by embedding patient-centered measures directly into clinical workflows [43]. Relatedly, future work should explore whether highly usable patient-facing systems could facilitate documenting pain-related goals and PEG scores, thereby reducing PCC burden and potentially increasing documentation rates even further than we observed among PCC OneSheet users.

For patients prescribed LTOT, encounters with OneSheet use were more likely to order UDS and review the PDMP compared with matched control encounters. The CDC guidelines recommend these practices as harm-reduction strategies, and PDMP review was also subject to state-level mandates in both study states during the study period, whereas UDS ordering was not [22–24]. This regulatory context likely contributed to the high baseline rate of PDMPD review (93%) observed prior to OneSheet implementation, limiting the magnitude of absolute improvement attributable to CDS system use. In contrast, ordering a UDS, a guideline-recommended practice not driven by state mandates, had lower baseline adherence and showed a larger absolute increase (28%) when OneSheet was used. Together, these findings suggest that CDS systems may be most impactful for improving guideline-recommended pain care in areas where baseline adherence is lower due to absence of regulatory requirements, rather than for practices already near universal compliance.

In contrast to the other outcomes, naloxone ordering was less common among encounters in which OneSheet was used. Although the absolute difference was small (1% versus 0.1%), this finding warrants cautious interpretation. During the study period, alternative naloxone distribution strategies (e.g., vending machine dispensing) and statewide standing orders reduced the need for clinician-issued naloxone prescriptions [44]. While these changes likely affected both treatment and control encounters, they may have reduced the extent to which clinician prescribing accurately reflected naloxone access. Additional research is needed to determine whether this finding reflects evolving naloxone distribution practices, residual confounding, or other unmeasured factors.

### Strengths and Limitations

This study has several strengths and limitations. We leverage data from an RCT combined with a per-protocol, propensity score matched approach, which reduces bias from observable patient, PCC, and encounter characteristics. By analyzing both chronic pain and LTOT encounters separately, we captured outcomes relevant to different clinical contexts. Several limitations should be considered when interpreting these findings. Encounters in which OneSheet was used likely represent a selected subset of clinicians, patients, and clinical situations. Although propensity score matching improved balance on measured characteristics, unmeasured factors associated with clinicians’ decisions to access OneSheet may remain. Consequently, findings should be interpreted as associations between OneSheet use and guideline-concordant care rather than estimates of the effect that would be expected under routine implementation across all eligible encounters. Lastly, several outcomes reflected guideline-concordant care status over predefined lookback period rather than clinical actions occurring exclusively during the indexed encounter. Consequently, some outcome events may have occurred before the recorded OneSheet access day. Findings should therefore be interpreted as associations between OneSheet use and guideline-concordant care rather than evidence that OneSheet directly caused specific clinical actions.

## Conclusion

Our per-protocol analysis suggests that OneSheet may support guideline-recommended chronic pain care when clinicians use the system. However, use was uncommon, limiting the potential population-level impact of the intervention. Future work should focus on identifying implementation strategies that promote routine use and workflow integration, as realizing the benefits of CDS systems may depend as much on successful implementation as on CDS functionality.

## Data Availability

The data used in this study contains sensitive patient information and are owned by participating health systems. Deidentified data are therefore not publicly available. Requests for access to study data may be directed to the Regenstrief Institute Data Services team at, subject to applicable data use agreements and institutional approvals.

## Acknowledgements

The authors acknowledge the contributions of the Regenstrief Institute’s Data Services (RDS) team, the clinical and information systems services teams at Eskenazi Health and Atrium Health Wake Forest Baptist Medical Center, and the primary care pain specialist teams at both health systems. Support from these teams was essential to OneSheet implementation and data collection.

## Notes

### Competing Interest Statement

The authors have declared no competing interest.

### Clinical Trial

NCT04295135

### Author Declarations

The Indiana University Institutional Review Board approved all study procedures before the study's commencement, including informed consent for participating PCCs and a waiver of consent for patients. Patient data was already routinely collected in the Epic EHR system, presenting no additional privacy risks for patients. Furthermore, the high volume of patients with chronic pain seen daily made obtaining individual consent and HIPAA authorization impractical.

### Funding

- National Institute on Drug Abuse: R33D1046085

